# Mask-aware foundational-model embeddings for 18F-FDG-PET/CT Prognosis in Multiple Myeloma

**DOI:** 10.1101/2025.11.04.25339482

**Authors:** Javier Guinea-Pérez, Silvia Uribe, Sara Peluso, Gastone Castellani, Cristina Nanni, Federico Álvarez

**Affiliations:** Universidad Politécnica de Madrid, Avenida Complutense, 30, Madrid, 28040, Madrid, Spain; Department of Medical and Surgical Sciences, Alma Mater Studiorum-University of Bologna, Via Massarenti, 9, Bologna, Italy; IRCCS Azienda Ospedaliero-Universitaria di Bologna, Bologna, Italy; Nuclear Medicine, IRCCS Azienda Ospedaliero-Universitaria di Bologna, Bologna, Italy

**Keywords:** Multiple Myeloma, Survival Analysis, Radiomics, Foundational models, Representation learning

## Abstract

**Purpose:** To test whether internal memory states from a medical foundational segmentation model can serve as compact, mask-aware embeddings for predicting progression-free survival (PFS) in multiple myeloma (MM) from whole-body [^18^F]FDG PET/CT, and how late fusion of PET, CT, and clinical data enhances prognostic performance.

**Methods:** We analyzed 227 newly diagnosed MM patients with PET/CT and clinical data. For two regions of interest (spine-dilated and full skeleton), we prompted MedSAM2 slice-wise using mask-derived bounding boxes and cached the final spatio-temporal memory tensor per modality. We compared two downsampling strategy to obtain per-study embeddings: channel×memory averaging with a small CNN head, and depth-attention pooling. PET and CT embeddings were combined by late fusion and passed to a DeepSurv head. We evaluated image-only and multimodal (image+clinical) models with stratified 5-fold cross-validation. The primary endpoint was Harrell’s c-index (mean ± SE across folds).

**Results:** Image-only models using the averaging downsampler achieved up to 0.659 ± 0.015 c-index (PET, spine-dilated), comparable to baseline radiomics results. Multimodal models improved discrimination to 0.710±0.032 (CT, spine-dilated), with similar performance for other PET/CT+clinical variants (0.703–0.710), improving clinical-only baselines ∼ 6.5%. Averaging consistently outperformed depth-attention; concatenation and gated fusion performed comparably. PET outperformed CT within the same mask in image-only settings.

**Conclusion:** Mask-aware memory embeddings extracted from a foundational segmentation model provide effective, data-efficient imaging biomarkers for MM PFS and, when fused with routine clinical covariates, significantly improve risk stratification over clinical-only or radiomics baselines. This offers a practical path to prognostic modeling on small medical cohorts without feature design.

## 1. Introduction

Multiple myeloma (MM) is a common cancer of bone marrow (BM) plasma cells that accounts for almost 10% of all hematologic diseases, having had more than 180000 diagnoses and 12000 deaths in 2022 alone [1, 2]. Few cases are diagnosed before metastasis (3% [3]), when disease markers are common and subtle symptoms [4]. Accurate risk stratification at diagnosis is central to tailoring therapy and surveillance, which can significantly improve 5-year survival rates [3].

Beyond clinical staging systems, imaging adds complementary information on tumor burden and disease biology. Whole-body [^18^F] fluorodeoxyglucose positron emission tomography-computed tomography (FDG PET/CT) has been included in the updated criteria for the diagnosis of MM of the International Myeloma Working Group [4] as it has proven to be useful for assessing the state of the disease and treatment strategies [5].

Risk stratification of patients is crucial for treatment selection [6], but it has not yet been delegated to imaging techniques. Standardized criteria for PET/CT interpretation in MM have been proposed [7], but visual interpretation can be difficult and remains limited in clinical practice [8]. Because of this, patient stratification is being carried out after invasive BM studies that identify chromosomal abnormalities [9]. FDG PET has the potential to improve risk stratification by detecting medullary and extramedullary focal lesions [10].

Recent years have seen a growing interest in the application of machine learning (ML) for FDG PET/CT interpretation in MM. Currently, images are processed using classical radiomic approaches, where handcrafted image features extract intensity, texture, and shape descriptors from segmented lesions. These features are then fed to ML algorithms with a clinical objective, be it risk stratification, biomarker discovery [11, 12], prognosis [13, 14], patient classification, diagnosis, and therapy response assessment [8]. These studies have shown that radiomic features can correlate with tumor burden and predict outcomes, but these features rely on manually predefined descriptors that can fail to capture high order interactions or spatial context across the skeleton.

In parallel, survival analysis has taken a step forward as deep learning-based survival models have extended traditional Cox proportional-hazards to model non linear survival data [15, 16]. These methods have been successfully integrated with radiomic feature extraction for survival analysis and biomarker discovery in MM and other ailments [17].

Foundational models are large-scale deep learning models trained on vast, heterogeneous datasets to acquire transferable representations that can be adapted to a wide range of downstream tasks with relatively little task-specific supervision [18]. In computer vision, examples include CLIP [19] and SAM [20], which have shown that training on diverse, high-volume datasets produces general-purpose embeddings that capture both semantic and structural image properties. These embeddings can then be fine-tuned or prompted to solve specialized tasks with far less annotated data than would otherwise be required. In the medical imaging domain, foundational models are being developed by pretraining on millions of scans spanning multiple modalities (CT, MRI, PET, ultrasound) and anatomical regions [21]. Adaptations of SAM, for example, have been fine-tuned to segment medical structures robustly across organ systems without retraining from scratch [22] [23].

In this work, we propose a survival analysis framework that overcomes the limitations of radiomics feature design by creating mask-aware data-efficient representations of FDG PET/CT volumes by using the internal memory state of a state of the art foundational segmentation model. These memory embeddings are generated during mask-guided volume propagation, where the model integrates anatomical prompts with imaging context. We extract and compress the final memory state into compact embeddings that capture information that radiomics descriptors cannot. We further explore late fusion strategies for PET and CT and evaluate our method in contrast to traditional radiomics features as well as measuring the improvement on a clinical feature only model.

In doing so, we address the gap between handcrafted radiomics and deep learning approaches: radiomics pipelines embed anatomical priors but are constrained by feature engineering, while deep survival networks can model complex non-linear risks but struggle to converge on small medical cohorts. Foundational medical imaging models provide a middle ground, offering robust, transferable representations that can be harnessed without retraining on small datasets. By leveraging the memory state embeddings of Med-SAM2, we show that it is possible to build clinically relevant, mask-aware survival predictors from FDG PET/CT in multiple myeloma. Related work has begun to explore SAM-derived embeddings for survival in 2D medical images in multi-omics datasets [24].

In summary, the contributions of this study are threefold: (i) we propose the use of foundational segmentation model memory states as a novel embedding for survival analysis; (ii) we benchmark PET, CT, and PET+CT late fusion embeddings against clinical and radiomics baselines; and (iii) we demonstrate that combining clinical variables with imaging embeddings yields superior progression-free survival (PFS) stratification. This positions foundational model embeddings as a promising bridge between radiomics and end-to-end deep learning in imaging-based prognostication for MM.

## 2. Materials and methods

### 2.1. Dataset

This study was based on an initial pool of 349 newly diagnosed MM patients collected since 2007 at IRCCS Azienda Ospedaliero–Universitaria Sant’Orsola-Malpighi (Bologna). All patients had baseline whole-body [^18^F]FDG PET/CT available. Four studies were excluded due to insufficient image quality and/or preprocessing failures that prevented reliable automatic mask generation (segmentation not obtainable), leaving 345 patients with usable imaging. From these, 262 patients had the required clinical annotations available (demographics/laboratory variables and outcome fields) and were retained for analysis; the remaining 83 were excluded due to missing clinical annotations. Finally, because PFS is the primary endpoint and therefore is not imputable, 35 patients without PFS time/event annotation were excluded, yielding a final analyzed cohort of 227 patients with PET/CT, clinical covariates, and PFS. A flow diagram summarizing these inclusion/exclusion steps and counts is provided in Figure 1.

**Figure 1:**
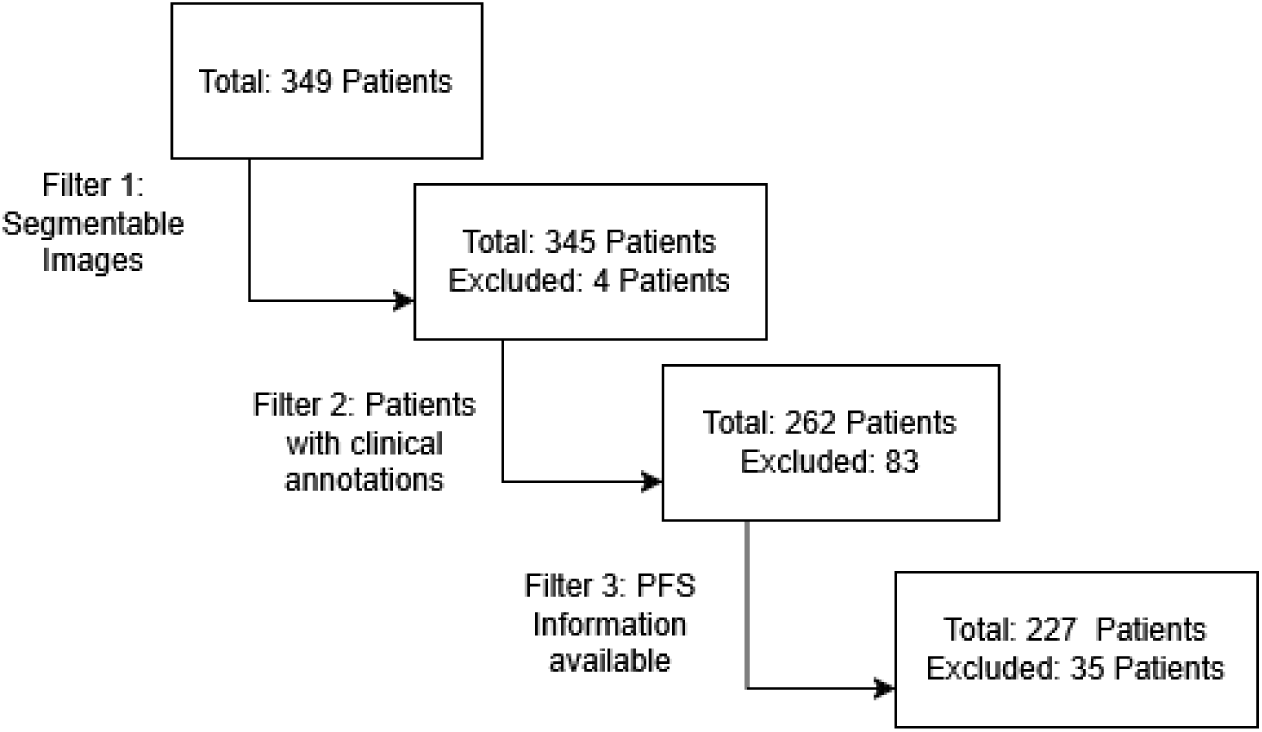
Exclusion criteria of available data.

This dataset has, to date, not been employed in the development of any public model, and therefore all external pretrained models had no patient-level overlap with the cohort.

PFS was defined as the time from diagnosis until relapse, progression, or death, with censoring applied to patients who did not experience an event during follow-up. The available dataset does not include event subtype labels (biochemical, radiologic, or clinical progression) or an explicit adjudication protocol; therefore, we modeled PFS as recorded.

Among 227 patients, 160 (70.5%) experienced a PFS event and 67 (29.5%) were censored. The observed PFS times had a median of 30 months (25.5 months among patients with events and 35 months among censored patients). Median follow-up, estimated using the reverse Kaplan–Meier method, was 51 months. The cohort included 132 men (58.1%) and 95 women (41.9%), with a mean age of 62.5 years (standard deviation 9.7 years). Baseline staging according to the Revised International Staging System (R-ISS) [25] was available for most patients: 49 (21.6%) were stage I, 95 (41.9%) stage II, 25 (11.0%) stage III, while staging information was not reported for 58 (25.6%) of the cases.

The dataset included clinical covariates with established relevance to MM prognosis: age, sex, creatinine, hemoglobin, platelets, antibody isotypes light chain ratio, and calcium. Additionally, the R-ISS stage (I/II/III) was present, which condensates information about clinical variables, some not present: *β*2 microglobulin, albumin, creatine, lactate dehydrogenase and chromosomal abnormalities. Variables with extensive missingness in this cohort (e.g., creatinine clearance) were excluded. Categorical variables were encoded and continuous variables standardized within each training fold prior to model fitting.

### 2.2. Preprocessing

PET and CT volumes were first loaded as NIfTI files and inspected. Because the two modalities differed in voxel origin and spacing despite identical qform/sform matrices, PET volumes were rigidly resampled onto the CT grid (same origin, voxel size, and dimensions) using linear interpolation with SimpleITK. No deformable registration or artifact correction was applied.

Mask generation followed the same automatic segmentation procedure described in our previous work [11]: CT scans were segmented using the Multi-Organ Objective Segmentation (MOOSE 2.0) model [26], which provides robust bone segmentation (median Dice coefficient 0.90 for most bones, except carpal, metacarpal and foot phalanges). From this segmentation, multiple regions of interest (ROIs) were derived through morphological operations aligned with the IMPeTUs protocol, which emphasizes bone marrow, spine, and paramedullary regions.

For the present study, we restricted the analysis to the two masks that demonstrated the best prognostic performance:

- **Spine dilated**: including vertebrae, internal spinal canal and surrounding paramedullary regions, designed to capture both intramedullary and paramedullary disease.
- **Skeleton**: a comprehensive mask encompassing the entire segmented skeleton and the dilated spine mask, designed to include total body bone disease burden.

Both masks were automatically generated on the CT images and then re-sampled to PET resolution, ensuring one-to-one alignment across modalities.

### 2.3. Architecture

Our pipeline converts FDG PET/CT volumes (paired with the automatically generated masks) into compact, mask-aware embeddings using the internal memory state of a foundational segmentation model, and then estimates the log-risk of progression with a DeepSurv head. PET and CT are processed in parallel with modality-specific embedding towers and are optionally combined with clinical data through late fusion before a survival head. Figure 2 summarizes the overall architecture.

**Figure 2:**
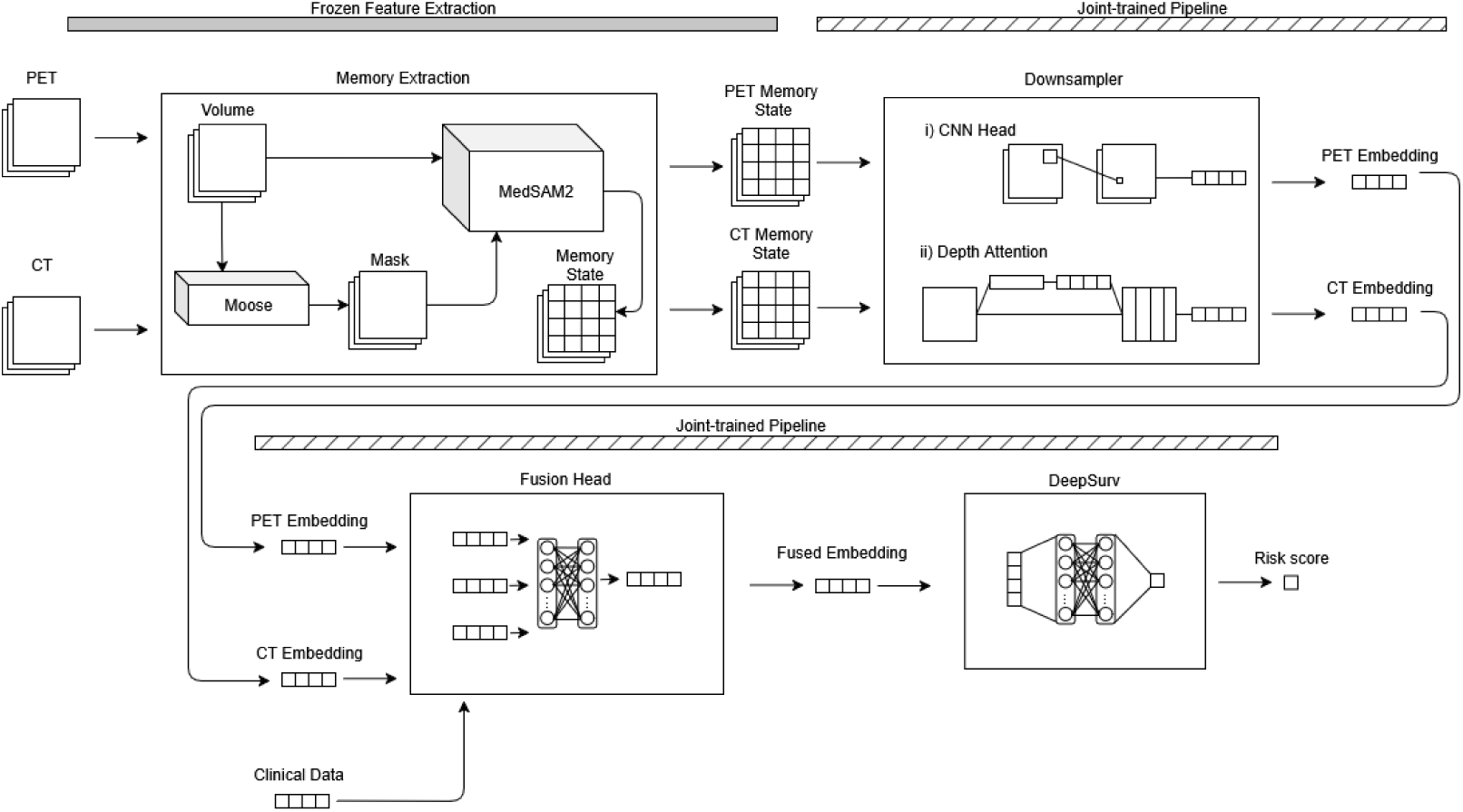
Overview of the proposed pipeline. The workflow is divided into four stages: (1) Memory Extraction: For each ROI (spine dilated or skeleton) and modality (PET or CT), axial slices are prompted with mask-derived bounding boxes and propagated through MedSAM2 [27]; the final memory state is cached. (2) Downsampler: The large memory tensor is compressed into a compact embedding (either with channel/memory averaging and a small CNN head and or a lightweight depth-attention block). (3) Fusion Head: Modality-specific embeddings are late-fused (with optional clinical covariates) into a single representation either using a learned gated sum or concatenation. (4) DeepSurv: A fully connected survival head outputs the linear predictor. The top progress bar indicates trainability. Memory Extraction is frozen while Downsampler, Fusion Head and DeepSurv are trained together end-to-end.

#### 2.3.1. Embedding generation

We extract image representations from **MedSAM2** [27], a medical adaptation of the SAM2 [28] segmentation foundation model. The model allows for segmentation of medical images with prompts by processing them as videos. In our case, for each study, we first select a ROI mask (either the *spine dilated* or the *full skeleton*, see Section 2.2). We generate prompts by computing the tight 2D bounding box of the ROI on each axial slice containing mask pixels and propagate the segmentation top-to-bottom through the volume. To alleviate memory requirements, only one of every two mask containing slices are propagated. MedSAM2 maintains a spatio-temporal memory during this slice-wise propagation; we cache the final memory state after the last slice and use it as our representation.

Bounding-box prompts were computed per axial slice as the minimal enclosing rectangle of the CT-derived ROI mask pixels. Prompting strategy was selected to maintain stable mask propagation through the volume and reasonable segmentation performance of the downstream MEDSAM2 model. This was verified by visual inspection on representative cases. During development we also tested alternative prompting modes (full-mask prompts and point prompts sampled within the ROI), which were less stable and/or yielded inferior downstream performance.

Concretely, for each modality (PET and CT) and ROI, MedSAM2 outputs a tensor of shape **M** ∈ R*^C^*^×^*^D^*^×^*^H^*^×^*^W^* (memory layers × channels × spatial height × width). In order to have all embeddings have the same memory layer size, they were cropped/padded in that dimension, ending with a tensor of shape: R^2×32×64×64^.

Given the cohort size, we consider two strategies to transform **M** into a compact embedding:

1. **Average embedding.** We perform global averaging across the memory and channel dimensions, reducing **M** to a 64×64 map. This 64×64 sized tensor was then processed through a small 2D convolutional neural network (CNN) head that maps an input with to a 128 × 1 sized embedding. Starting from (1, 64, 64), the two 2 × 2 max-pool layers reduce the resolution to (64, 16, 16). A final 3 × 3 convolution produces (128, 16, 16), which is collapsed by global average pooling (GAP, implemented as AdaptiveAvgPool2d(1, 1)) to (128, 1, 1) and flattened to a vector in R^128^. This head is intentionally small to limit capacity and overfitting given cohort size, while GAP provides translation-robust, size-agnostic summarization of local patterns learned by the convolutions. Figure 3a details the process.
2. **Light attention embedding.** Given a memory tensor *x* ∈ R*^C^*^×^*^D^*^×^*^H^*^×^*^W^*, we first reduce the in–plane resolution with average 3D pooling over (1, 2, 2) to obtain (*C, D,* 32, 32). A 1×1×1 pointwise 3D convolution mixes the *C* input channels into a compact width *base*, followed by normalization and GELU, yielding (*base, D,* 32, 32).

**Figure 3:**
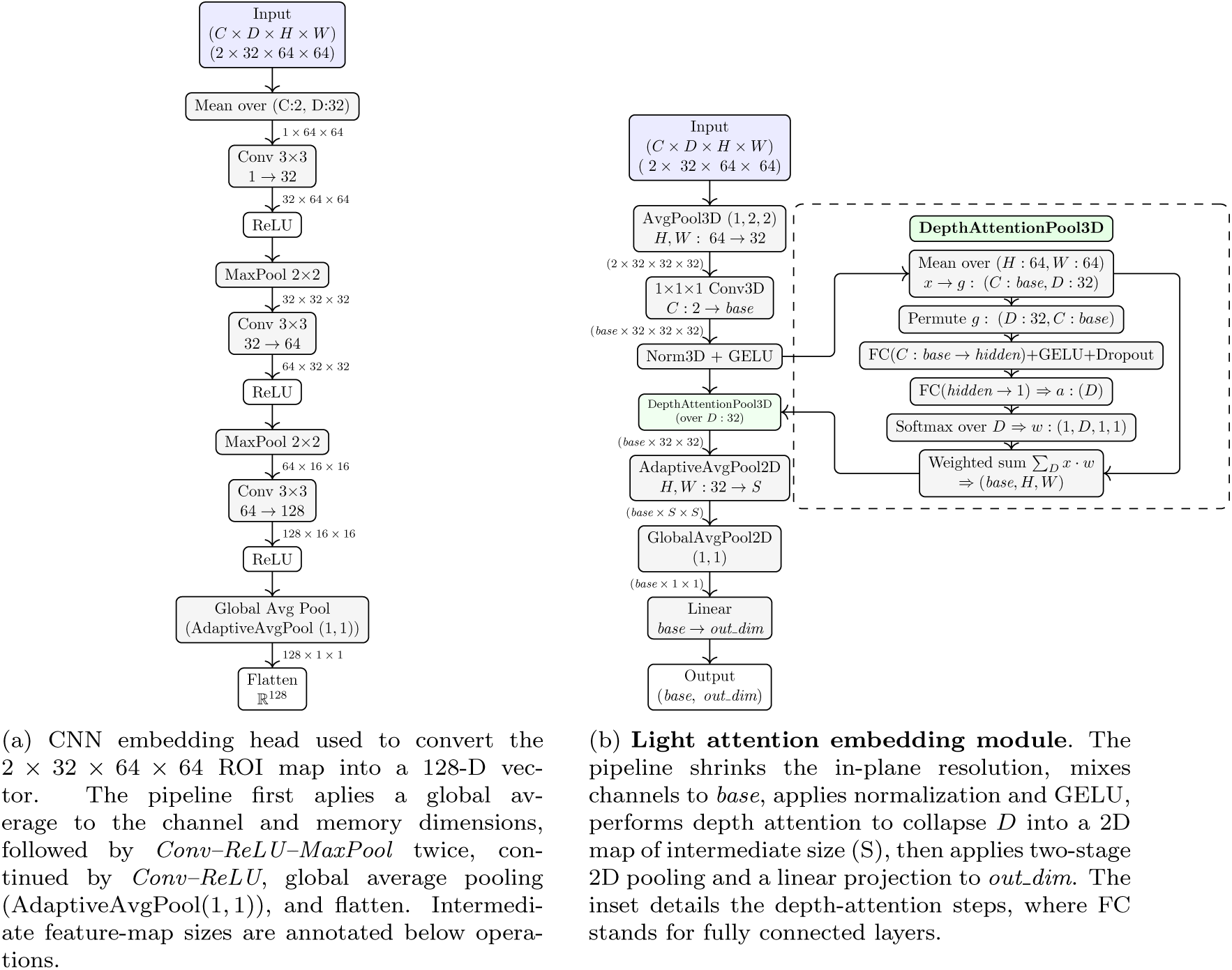
Downsampler architectures: (a) average embedding head; (b) light attention head.

Our deph attention block follows the squeeze-and-excitation operator [29]. We squeeze spatial information by global average pooling over (*H, W*) to produce depth descriptors *g* ∈ R*^base^*^×^*^D^*. A two-layer MLP

with GELU then excites these descriptors to obtain slice-wise logits, and a softmax across *D* provides normalized attention weights *w* ∈ R^1×^*^D^*. The weighted sum collapses depth:

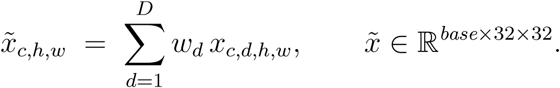

The resulting 2D map is resized to *S*×*S* via AdaptiveAvgPool2D, globally averaged to (*base,* 1, 1), flattened to R*^base^*, and linearly projected to the final embedding ∈ R*^out^ ^dim^*. Figure 3b details the attention pipeline.

*Discarded configurations.* We also evaluated three additional downsampling heads: (i) a 3D CNN head operating on ((*C*×*D*×)64×64), (ii) direct flattening of the 64×64 maps, and (iii) PCA-based flattening, all under the same outer splits and training protocol. All three variants performed at chance (mean c-index ≈ 0.50, high p-values) and were therefore excluded from the final pipeline.

#### 2.3.2. Fusion head

We combine modality embeddings (and optionally clinical covariates) via late fusion before the survival head.

*(i) Concatenation (1D stacking)..* Each modality-specific tower outputs a fixed-length vector **z**_PET_, **z**_CT_ ∈ R*^d^*. Concatenation is performed as simple 1D stacking:

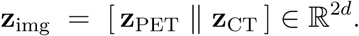

When including clinical variables, a vector **z**_clin_ ∈ R*^d^*^clin^, the final fused input to the survival head is:

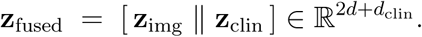

This scheme is parameter-efficient, preserves modality-specific information, and naturally supports single-modality inputs.

*(ii) Lightweight gated fusion (scalar mixing)..* Alternatively, we add modality vectors **z**_PET_, **z**_CT_ ∈ R*^d^* using a scalar gate *α* ∈ (0, 1) learned via logistic regression on their 1D concatenation:

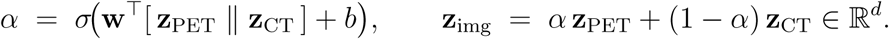

This gate adds only 2*d* + 1 parameters and yields an interpretable per-case modality weighting.

If clinical covariates are used, we again stack 1D:

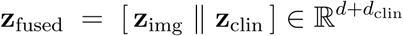

#### 2.3.3. DeepSurv Head

The survival head maps the fused vector **z**_fused_ to a scalar linear predictor 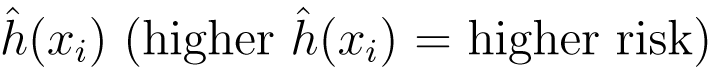 using a deep feed-forward neural network. We update the standard DeepSurv model [15] to include weight decay, and early stopping on validation partial likelihood. The model is trained with the average negative log partial likelihood with regularization as the loss function:

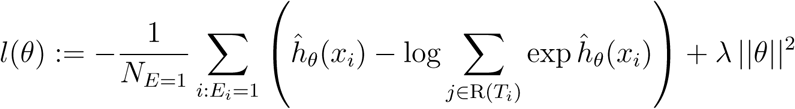

where *N_E_*_=1_ is the number of observed events, *θ* the parameters of the survival head, R(*T_i_*) is the risk set just prior to *T_i_*, and *λ* controls weight decay. Dropout and batch normalization are applied to hidden layers.

### 2.4. Experiments

#### 2.4.1. Experimental setup

We evaluated three families of models on PFS: (i) image-only embeddings from PET/CT, (ii) *clinical-only* baselines, and (iii) *multimodal* models combining imaging embeddings with clinical covariates. MedSAM2 memory extraction was frozen, while the downsampler, fusion, and DeepSurv head were trained end-to-end per training fold. We used stratified 5-fold cross-validation preserving event proportion and approximate time quantiles. Optimization used AdamW cosine/step decay, dropout, BN, gradient clipping, and early stopping on validation partial likelihood. Hyperparameters were tuned by maximizing 5-fold cross-validated concordance using Sobol sequences in Optunity [30] to search the parameter space. Because of the limited cohort size and computational cost of nested hyperparameter optimization, we used an internally tuned cross-validation procedure with a restricted search space and strong regularization. This may yield mildly optimistic performance estimates [31]. Detailed search ranges and results are provided in the supplementary spreadsheet.

#### 2.4.2. Models setup

*Image-only.* We ablated (a) modality: PET, CT, PET+CT; (b) ROI: Spine-dilated vs Skeleton; and (c) downsampler: Average embedding vs Light attention. PET+CT used late fusion (Section 2.3.2) and additionally tested fusion strategies: concatenation vs gated mixing. We also compare these configurations against a baseline non-medical image embedding model, ablating this way the fine-tuning and mask components of the pipeline.

*Clinical-only.* Two clinical baselines were trained using the dataset’s clinical variables (Section 2.1): CoxPH (ridge-penalized) and DeepSurv. Categorical variables were one-hot encoded, continuous variables were *z*-scored within each training fold and missing values were imputed from training-fold statistics.

*Multimodal.* We extend the best image-only configuration (downsampler + fusion) by adding clinical covariates. We report PET+CT+Clinical and single-modality+Clinical variants to assess complementarity with imaging. The same clinical features and preprocessing as in the clinical only model were used.

#### 2.4.3. Metrics and statistical analysis

Our primary endpoint is the mean Harrell’s c-index [32] on outer test folds, reported as mean ± standard error (SE). For the best performing model we include the Kaplan-Mayer curve of the least and most at risk patient subsets. Significance is assessed with a one-sample *t*-test against a random model (c-index of 0.5); model comparisons use paired *t*-tests on fold-wise differences (as we used identical splits).

## 3. Results

We evaluated image-only ablations, multimodal variants, and baselines. Summary metrics are in Tables 1–3.

**Table 1:**
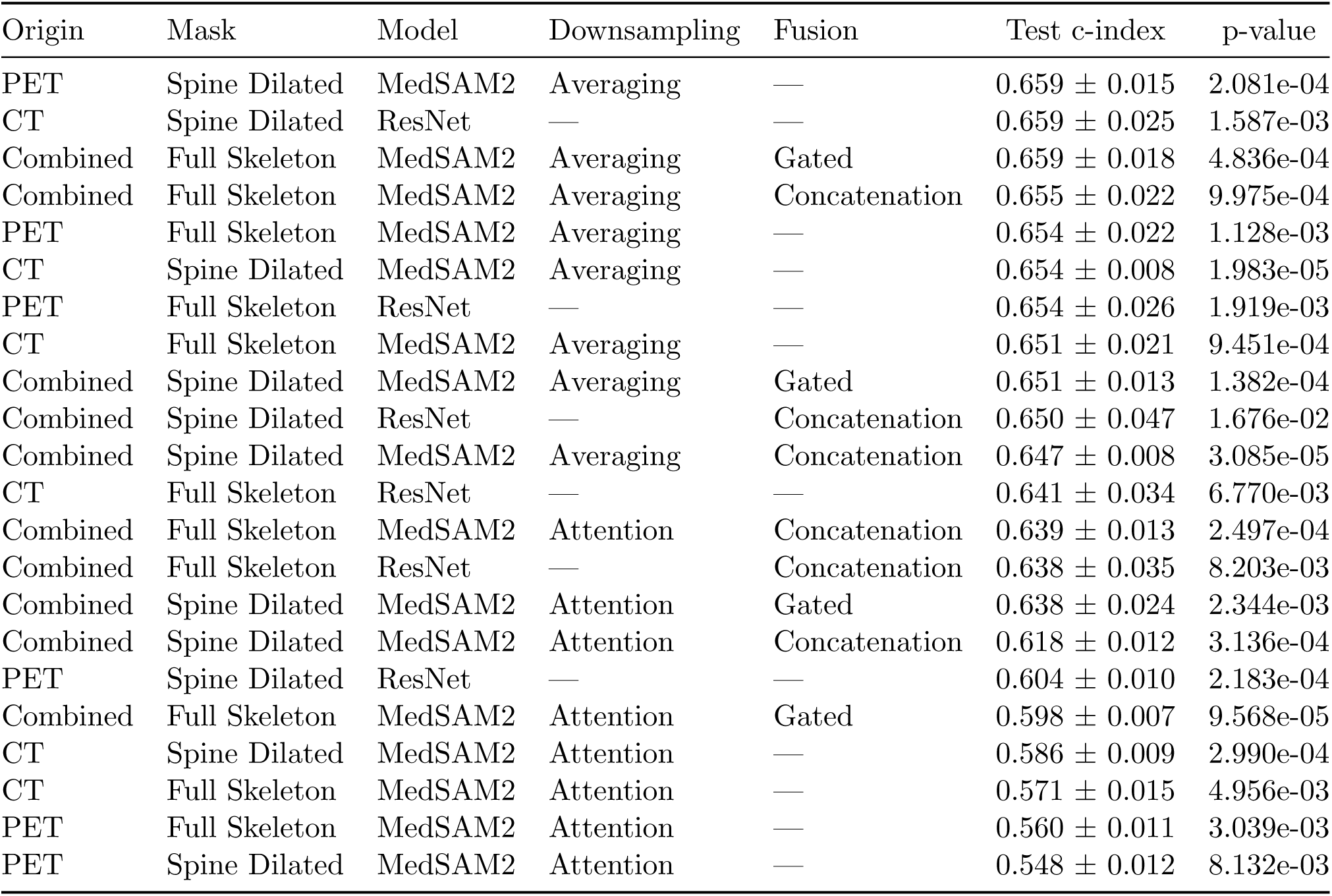
Image-only configurations (sorted by c-index). Reported c-indexes are mean ± SE. An en dash (—) denotes not applicable.

**Table 2:**
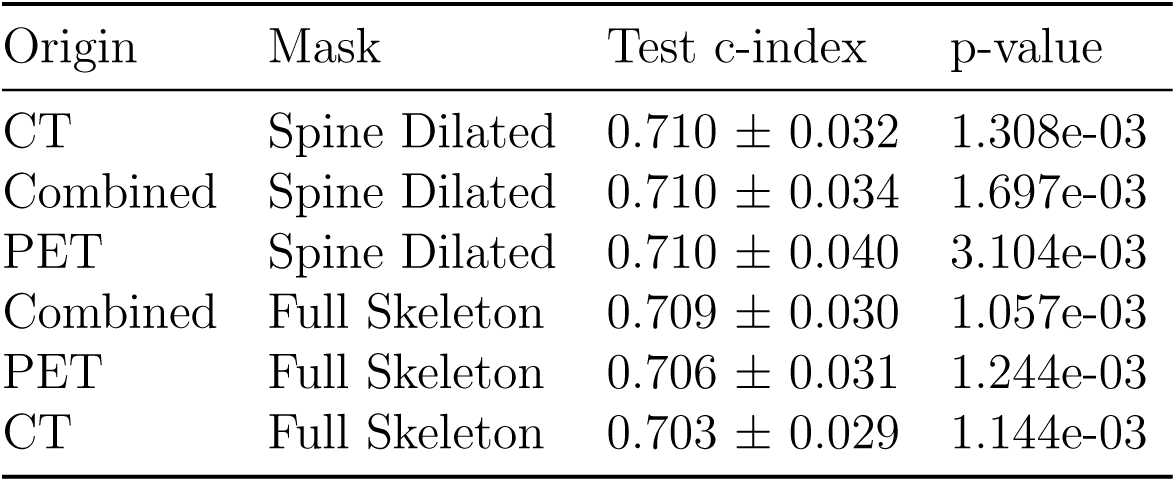
Multimodal configurations (sorted by c-index).

**Table 3:** Best models c-index (5-fold cross validation mean ± SE).

| Model | c-index |
| --- | --- |
| Multimodal Model - Our Embedding + Clinical | $0.710 \pm 0.030$ |
| Clinical Only - DeepSurv | $0.667 \pm 0.035$ |
| Clinical Only - Cox | $0.661 \pm 0.025$ |
| Image Only - Our Embedding | $0.659 \pm 0.014$ |
| Image Only - Radiomics Baseline | $0.657 \pm 0.016$ |

Ablations on downsampling and fusion were conducted using image-only models (Table 1). Across origins and masks, averaging consistently out-performed attention. The top results were obtained with (i) PET + Spine Dilated + Averaging (no image fusion; 0.659 ± 0.015, *p* = 2.08×10^−4^) and (ii) Combined + Full Skeleton + Averaging with Gated fusion (0.659 ± 0.018, *p* = 4.84×10^−4^).In contrast, Attention-based variants underperformed across all configurations (0.548–0.639 c-index).

The embedding pre-trained CNN baseline (ResNet [33]) achieved competitive performance in some image-only settings (Table 1), including one of the top configurations, but showed higher fold-to-fold variability (larger SE) than MedSAM2 memory embeddings. Across most configurations, Med-SAM2 provided comparable or better discrimination with more consistent performance.

Multimodal configurations were trained using the best downsampling choice from the ablation, that is, averaging, and are summarized in Table 2. The best multimodal configuration reached 0.710 ± 0.032 (CT + Spine Dilated), closely followed by Combined + Spine Dilated (0.710 ± 0.034) and PET + Spine Dilated (0.710 ± 0.040). Compared with the best image-only result, this corresponds to a relative improvement of ∼ 7.8%; compared with the best clinical baseline (Table 3), the improvement is ∼ 6.5%.

Figure 4 shows Kaplan–Meier curves for the first and fourth risk quartiles produced by the best overall model (clinical + CT with the spine mask). Quartiles were computed from out-of-fold risk estimates standardized using in-fold statistics. The high-vs low-risk separation is statistically significant (log-rank *p* = 3.14×10^−3^).

**Figure 4:**
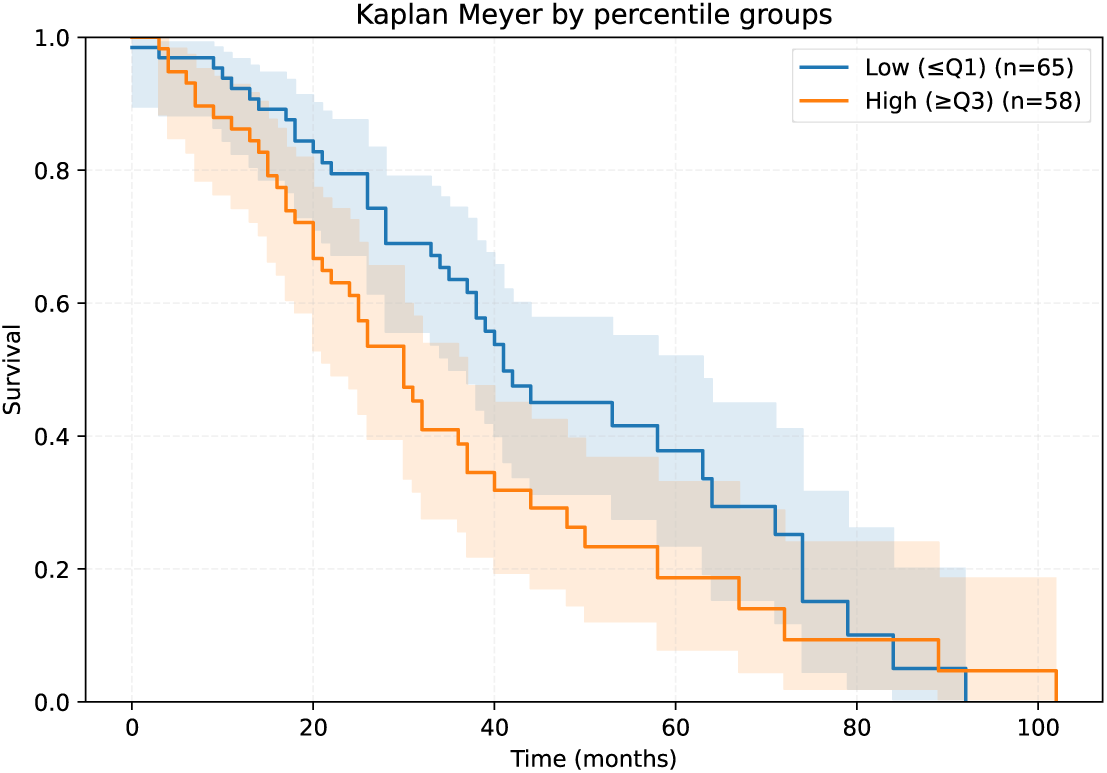
Kaplan-Meier curves of the first and fourth quartile risk patients from the best performing model (clinical variables + CT with the spine mask). Risk quartiles calculated from out-of-fold risk estimations standardized with in-fold estimations. Log-rank test p-value of 3.14e-03.

## 4. Discussion

Our novel embedding strategy proves the viability of the usage internal states of foundational medical segmentation models as information rich embeddings for survival analysis in MM. Not only are the results comparable with radiomics models (without defining the features), but results show that our embedding added to clinical models shows an improvement to clinical only models (see Table 3).

In MM, prognostic stratification can influence the overall intensity of therapy, transplant planning, and the intensity of maintenance and surveillance [34]. Consequently, the risk score produced by our model is best viewed as an imaging-derived adjunct biomarker that could complement established clinical and laboratory stratification rather than replace it. Given the retrospective single-center design, limited cohort size, and incomplete availability of some guideline-grade prognostic variables in this dataset, we do not propose clinical decision thresholds or treatment changes based on our predictions; these steps require calibration and external multi-center validation with threshold-dependent utility analyses, as is explained in more detail in Section 5.

The comparison with the ResNet baseline indicates that generic visual embeddings can recover some prognostic signal from PET/CT, even without explicit segmentation prompting. However, MedSAM2 memory embeddings are explicitly conditioned on ROI prompts and integrate information accumulated during mask-guided propagation, which may act as an inductive bias aligned with disease-relevant anatomy. In our experiments, this translated into more consistent performance across folds (lower SE) and competitive or superior c-index relative to ResNet. These findings support the view that mask-aware memory states provide a practical middle ground between hand-crafted radiomics and generic pre-trained encoders when cohort sizes are limited.

In all settings (modality and mask), simple averaging across memory layers and channels outperformed the light attention downsampling alternative (Figure 6 shows the difference between downsampling strategies). Memory states are highly redundant and smooth across slices, so averaging behaves as a variance-reducing low-pass filter that suppresses slice-specific noise and propagation artifacts. Additionally, the attention head introduces additional degrees of freedom that might not be reliably constrained, increasing the risk of overfitting. Consistent with this interpretation, averaging yielded lower standard errors and p-values.

With respect to multi-image setups, fusion of PET and CT, Fig. 5 shows that 1D concatenation and scalar-gated mixing deliver statistically indistinguishable discrimination (paired tests, *p >* 0.05). Both strategies access the same modality-specific vectors, and the DeepSurv head can effectively reweight concatenated features. Nevertheless, the gate remains attractive from an interpretability standpoint, as it provides a per-patient estimate of modality importance.

**Figure 5:**
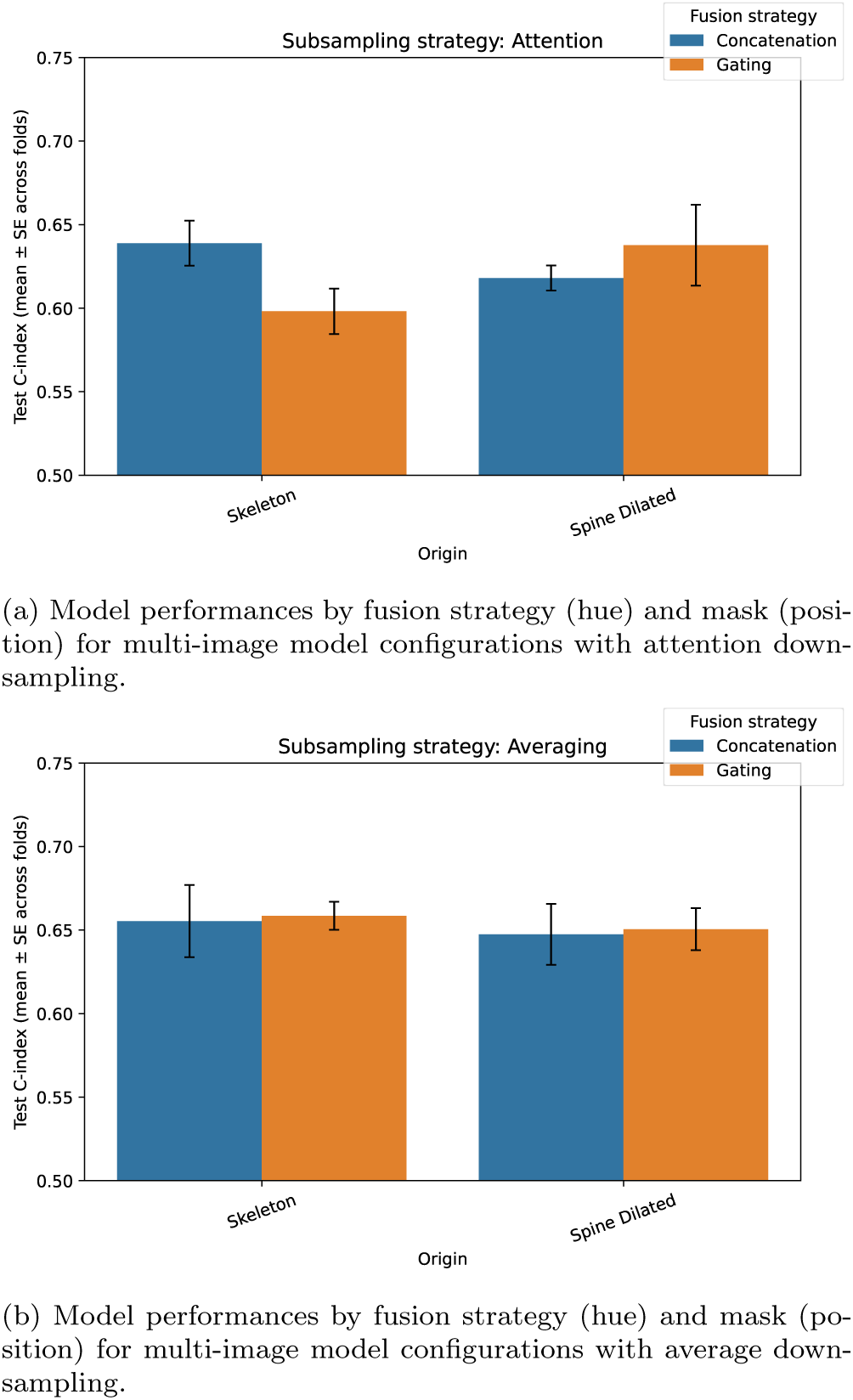
Comparison between both fusion strategies in imaging only models.

**Figure 6:**
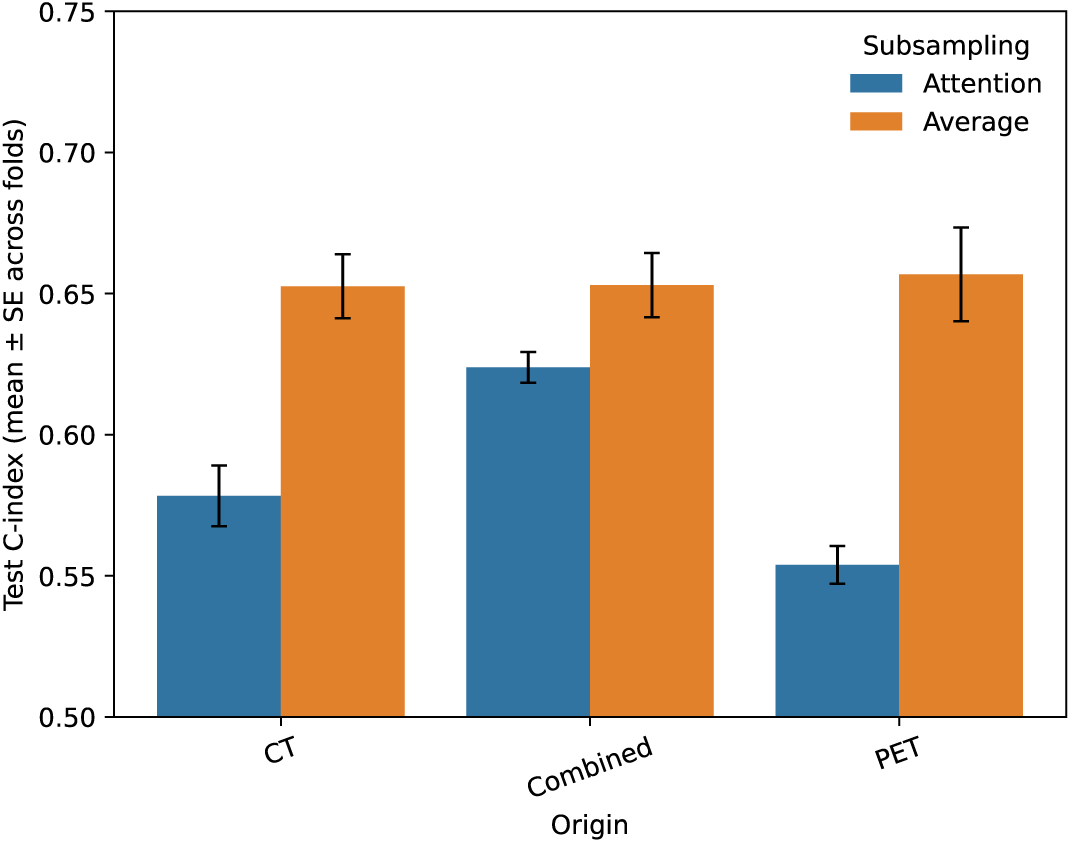
Aggregated model performances by downsampling strategy (hue) and origin (position) for multi-image model configurations. Reported c-index values correspond to the mean and error across folds, obtained by first averaging the c-index of each model within each fold and then aggregating these fold-wise means across the cross-validation splits.

Within image-only models, PET consistently outperformed CT under the same mask, in line with prior work and clinical research [11, 35].

Finally, mask differences were statistically insignificant both in unimodal and multimodal models (p-values *>* 0.05).

## 5. Limitations and future work

Limitations of this work include a small dataset with limited clinical information. The number of events restricts statistical power, increases fold-to-fold variability, and constrains the capacity of the downsampler and fusion heads. Detailed acquisition and reconstruction metadata (scanner model, PET reconstruction settings, injected dose/uptake time, and CT parameters) were not available in this retrospective cohort, preventing explicit adjustment for scanner/protocol effects and leaving open the possibility of residual acquisition-related confounding. Additional, external, multi-center validation is needed to assess generalization under scanner/reconstruction variability. External validation would also allow for a less biased assessment, as internally tuned estimates may be mildly optimistic. Also, due to missing clinical values being imputed and becuase the missingness mechanism is unknown, residual bias in clinical-only and fusion models cannot be excluded. Transferability may be limited by differences in upstream preprocessing and ROI-mask generation across sites, since our prompting is mask-driven and segmentation errors can shift the resulting memory embedding distribution. Consequently, models and hyperparameters tuned on our cohort may not be optimal under a new center’s data distribution and should be re-tuned during external validation.

Residual PET/CT misregistration (for example, due to motion or breathing) was not explicitly quantified in this study. Even though ablations show that discrimination performance is similar between unimodal sources; future work could incorporate modality alignment metrics and assess robustness to registration error.

The underperformance of the proposed lightweight attention downsampler should not be interpreted as evidence against attention mechanisms in general; higher-capacity attention architectures may behave differently but were not explored here due to sample-size constraints. With a large enough dataset, both this method or other attention mechanisms might perform better.

The retrospective dataset provides PFS as a curated time-to-event end-point but does not include granular progression subtype labels or a documented adjudication protocol; future work using standardized event subtypes could reduce endpoint heterogeneity and enable more clinically specific modeling.

Additionally, the clinical covariates used when building the multimodal model as well as the clinical only baseline were limited. We only used available variables with acceptable completeness. Using richer multi-omics variables would likely improve performance and allow for a deeper explainability analysis.

Subgroup analyses (e.g., transplant eligibility, cytogenetic risk, extramedullary disease) were not performed because these annotations were unavailable or incomplete and subgroup sizes would be underpowered; additional robustness testing under alternative split schemes and external validation remain future work.

Our mask prompts introduce a prior over “where to look”: masks focus the model on regions deemed anatomically informative. Although prompts were derived from an automatic CT segmentation trained independently of the folds, residual concerns about prompt-induced bias remain, as well as how much of the signal comes from the memory versus the prompt prior. Future work could compare our mask-derived prompts to coarser boxes and random prompts; and evaluate whether prompting on anatomically uninformative regions degrades performance to chance, clarifying the contribution of mask priors.

We did not perform a formal quantitative sensitivity analysis of embedding stability under prompt perturbations (bounding-box jitter/padding changes or mask boundary noise), which remains important future work to further validate robustness of the memory embeddings.

Our baseline comparisons currently include a standard pre-trained CNN (ResNet) but not transformer-based encoders (e.g., ViT) or the fine tuned image-encoder component of SAM2 (Hiera [36]) in isolation. Future work should benchmark these alternatives and explicitly disentangle the contribution of the SAM2-family encoder versus prompt-guided propagation and memory updates.

The scalar-gated fusion provides per-patient modality weighting that may aid case review; projecting embedding saliency back to the masks (e.g., Grad-CAM-style analysis over the downsampler) could further expose image patterns associated with risk, a deeper explainability study could explore these ideas. In Appendix A we show an example of anatomical interpretability, though because in this pipeline there is no differentiable path between the risk prediction and the image, we have opted for exploring occlusion sensitivity.

## 6. Conclusion

This study demonstrates that mask-aware memory embeddings derived from the internal states of a foundational segmentation model can serve as effective imaging biomarkers for PFS in multiple myeloma.

Without handcrafting features, our memory-based embeddings achieved performance comparable to a strong radiomics baseline and improved a clinical-only model when fused with routine covariates (Table 3). These results support the central premise that foundational medical models encode rich, transferable information that can be repurposed—via lightweight heads and rigorous evaluation—to address prognostic tasks in small, censored cohorts.

Simple averaging across memory×channel dimensions consistently out-performs a lightweight attention alternative, suggesting a favorable bias–variance trade-off in this data regime. PET outperforms CT under identical masks, consistent with metabolic burden driving risk, while late-fusion choices (concatenation vs. scalar gating) deliver comparable discrimination.

Overall, foundational-model memory embeddings offer a bridge between handcrafted radiomics and end-to-end deep survival networks: anatomically grounded, computationally light, and readily extensible to multimodal fusion.

## Appendix A. Anatomical interpretability via occlusion sensitivity

The proposed pipeline uses MedSAM2 as a frozen embedding generator: for each study, PET/CT volumes are prompted slice-wise with mask-derived bounding boxes, MedSAM2 propagates segmentation information through the volume, and the final internal memory state is cached and compressed into a compact representation. Because the embedding extractor is not trained end-to-end and embeddings are stored offline, standard gradient-based explainability methods like GradCAM are not directly applicable to the full imaging-to-risk mapping as implemented. To obtain spatially grounded explanations under these constraints, we adopt a perturbation-based occlusion sensitivity analysis that quantifies how localized image perturbations alter the predicted risk.

Given a trained survival model (downsampler + fusion + DeepSurv) and a frozen MedSAM2 embedding extractor, we estimate an anatomical importance map by repeatedly occluding small regions of the input image, regenerating the corresponding memory embedding under the same prompting protocol, and measuring the resulting change in the model’s predicted log-risk. Regions whose occlusion causes the largest change in predicted risk are interpreted as most influential for the model’s prediction, conditional on the prompted anatomical ROI.

For each selected case, we perform the following steps:

1. **Baseline embedding and risk.** Generate the MedSAM2 memory embedding using the standard prompting protocol (slice-wise tight bounding boxes derived from the ROI mask) and compute the baseline log-risk with the trained DeepSurv head.
2. **Define an occlusion grid.** Inside the ROI, define a regular grid of patch locations. To keep the analysis anatomically constrained, we only evaluate patches whose centers lie inside the ROI mask.
3. **Occlude and re-embed.** For each patch location, modify the image by replacing voxel intensities inside the patch with a constant value (we use a neutral fill value, the within-volume mean). We then re-run MedSAM2 propagation using the same bounding-box prompts and cache the resulting memory embedding.
4. **Risk delta.** Pass the occluded embedding through the same downsampler/fusion/DeepSurv head to obtain a new log-risk. The difference to the baseline log-risk (signed or absolute) is recorded as the patch im-portance score.
5. **Build an importance map.** Assign each patch score back to the occluded spatial region and aggregate overlapping patches (by averaging) to obtain a smooth voxel-level importance map. We visualize this map as overlays on representative slices or projections within the ROI.

This analysis shows which anatomical regions, when disrupted, most change the model’s predicted risk. Because it is perturbation-based, it provides a model-faithful sensitivity measure for the complete pipeline. It does not imply causality, and the resulting maps depend heavily on design choices such as patch size, stride, and occlusion fill value; therefore, we interpret the visualizations qualitatively as indicators of spatial sensitivity.

Due to data-sharing restrictions for our institutional MM cohort, we cannot publish patient-level overlays from the training dataset. We therefore include an illustrative example (Figure A.7) computed on a representative public whole-body PET/CT case (not used in model development and not diagnosed with MM) to demonstrate the procedure and verify that the resulting sensitivity patterns are spatially localized within anatomically plausible parts of the prompted ROI. The image has been sourced from The Cancer Imaging Archive (TCIA) image collection [37], and the sensitivity analysis was performed in the best performing image only model. Future work should extend this analysis to cohort-level aggregated attribution statistics under appropriate governance.

**Figure A.7:**
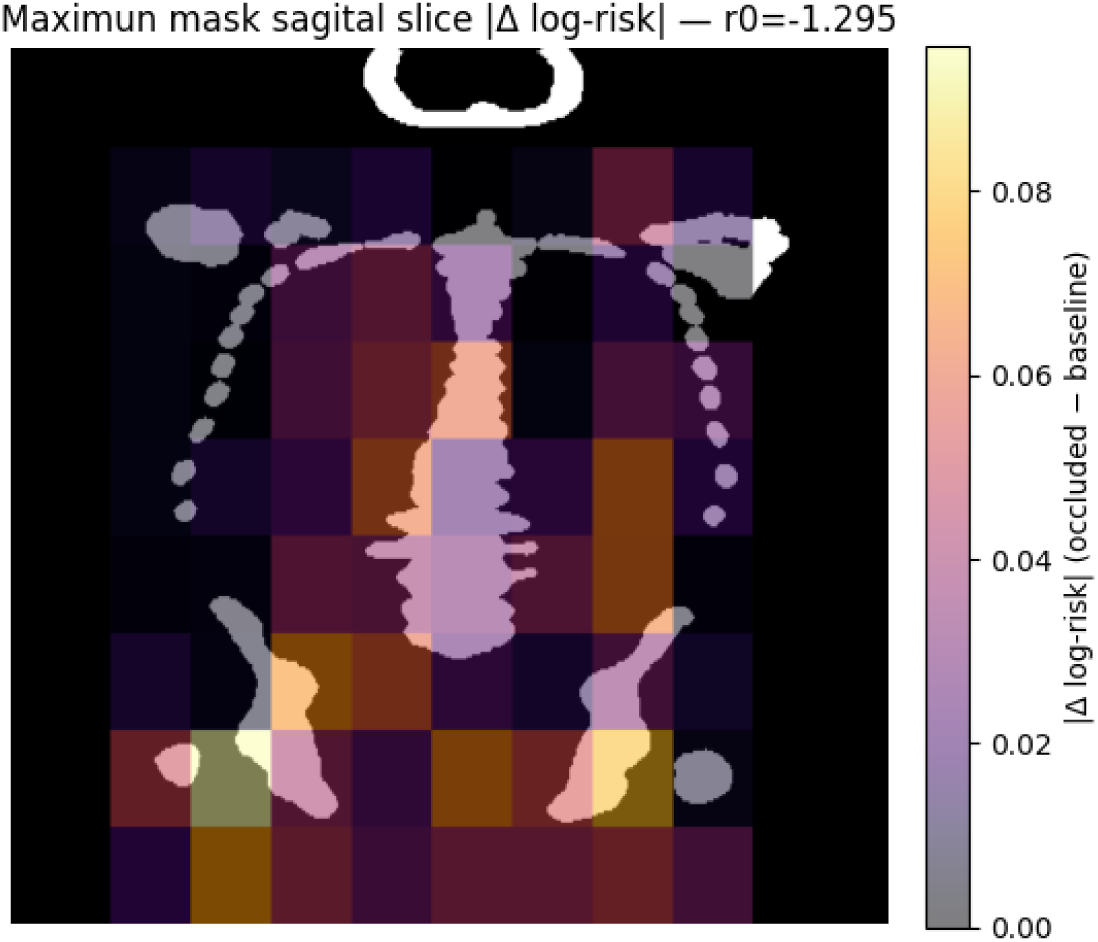
Occlusion sensitivity visualization (anatomical interpretability). Heatmap of risk sensitivity obtained by patch-wise occlusion within the prompted skeletal ROI and recomputation of the predicted log-risk. The heatmap is overlaid on the sagittal slice with the largest ROI area for a representative public whole-body PET/CT example (not used for model training). Warmer colors indicate regions where occlusion causes a larger change in predicted log-risk. In this case, the highest sensitivity is concentrated in the hip and proximal lower-limb regions within the ROI.

## Data Availability

All data in the present study is private.

## Supplementary information

Spreadsheet with optimization range and best model hyperparameters available.

All code required to reproduce the proposed embedding extraction, training, and evaluation pipeline is available at https://gitlab.com/synthiaeu/memory-embeddings. The repository includes implementation and configuration files for MedSAM2 memory extraction, downsampling heads, late fusion, DeepSurv training, and cross-validation evaluation, occlussion sensi-tivity analysis, along with documentation for integrating new datasets.

## Acknowledgements

Thanks to Matteo Bastico for the idea and to Pablo Perdomo-Quinteiro, Alessandro Ceresi, Anaida Fernández Garćıa and Francisco Moreno Garćıa for their comments and support.

## CRediT authorship contribution statement

**Javier Guinea-Pérez**: Conceptualization, Methodology, Software, Formal Analysis, Investigation, Writing - Original Draft, Writing - Review & Editing, Visualization. **Silvia Uribe**: Writing - Review & Editing, Funding acquisition. **Sara Peluso**: Writing - Review & Editing, Data Curation, Resources. **Gastone Castelani**: Resources, Funding acquisition, Project administration.**Cristina Nanni**: Resources, Data Curation. **Fed**erico **Álvarez**: Supervision, Writing - Review & Editing, Methodology, Conceptualization, Funding acquisition, Project administration.

## Ethics statement

The data used in this work was gathered by IRCCS Azienda Ospedaliero-Universitaria Sant’Orsola-Malpighi in Bologna in compliance with Italian law.

Data was accessed by UPM researchers with ethics compliance following Genomed (Grant no. 101017549) and SynthIA’s (Grant No. 101172872) data sharing agreements.

## References

[1] A. Mafra, M. Laversanne, R. Marcos-Gragera, H. V. S. Chaves, C. Mcshane, F. Bray, A. Znaor, The global multiple myeloma incidence and mortality burden in 2022 and predictions for 2045, JNCI: Journal of the National Cancer Institute (2024) djae321doi:10.1093/jnci/djae321.

[2] H. Ludwig, S. Novis Durie, A. Meckl, A. Hinke, B. Durie, Multi-ple Myeloma Incidence and Mortality Around the Globe; Interrelations Between Health Access and Quality, Economic Resources, and Patient Empowerment, The Oncologist 25 (9) (2020) e1406–e1413. doi:10.1634/theoncologist.2020-0141.

[3] SEER*Explorer Application. URL https://seer.cancer.gov/statistics-network/explorer/application.html?site=1&data_type=1&graph_type=2&compareBy=sex&chk_sex_3=3&chk_sex_2=2&rate_type=2&race=1&age_range=1&hdn_stage=101&advopt_precision=1&advopt_show_ci=on&hdn_view=0&advopt_show_apc=on&advopt_display=2#resultsRegion0

[4] S. V. Rajkumar, M. A. Dimopoulos, A. Palumbo, J. Blade, G. Merlini, M.-V. Mateos, S. Kumar, J. Hillengass, E. Kastritis, P. Richardson, O. Landgren, B. Paiva, A. Dispenzieri, B. Weiss, X. LeLeu, S. Zweegman, S. Lonial, L. Rosinol, E. Zamagni, S. Jagannath, O. Sezer, S. Y. Kristinsson, J. Caers, S. Z. Usmani, J. J. Lahuerta, H. E. Johnsen, M. Beksac, M. Cavo, H. Goldschmidt, E. Terpos, R. A. Kyle, K. C. Anderson, B. G. M. Durie, J. F. S. Miguel, Interna-tional Myeloma Working Group updated criteria for the diagnosis of multiple myeloma, The Lancet Oncology 15 (12) (2014) e538–e548. doi:10.1016/S1470-2045(14)70442-5. URL https://www.thelancet.com/article/S1470-2045(14)70442-5/fulltext

[5] A. Agarwal, A. Chirindel, B. A. Shah, R. M. Subramaniam, Evolving role of FDG PET/CT in multiple myeloma imaging and management, AJR. American journal of roentgenology 200 (4) (2013) 884–890. doi: 10.2214/AJR.12.9653.

[6] S. V. Rajkumar, S. Kumar, Multiple myeloma current treatment al-gorithms, Blood Cancer Journal 10 (9) (2020) 94. doi:10.1038/s41408-020-00359-2. URL https://www.nature.com/articles/s41408-020-00359-2

[7] C. Nanni, A. Versari, S. Chauvie, E. Bertone, A. Bianchi, M. Rensi, M. Bellò, A. Gallamini, F. Patriarca, F. Gay, B. Gamberi, P. Ghedini, M. Cavo, S. Fanti, E. Zamagni, Interpretation criteria for FDG PET/CT in multiple myeloma (IMPeTUs): final results. IMPeTUs (Italian myeloma criteria for PET USe), European Journal of Nuclear Medicine and Molecular Imaging 45 (5) (2018) 712–719. doi: 10.1007/s00259-017-3909-8.

[8] L. Manco, D. Albano, L. Urso, M. Arnaboldi, M. Castellani, L. Florimonte, G. Guidi, A. Turra, A. Castello, S. Panareo, Positron Emission Tomography-Derived Radiomics and Artificial Intelligence in Multiple Myeloma: State-of-the-Art, Journal of Clinical Medicine 12 (24) (2023) 7669. doi:10.3390/jcm12247669. URL https://www.mdpi.com/2077-0383/12/24/7669

[9] S. K. Kumar, S. V. Rajkumar, The multiple myelomas — current concepts in cytogenetic classification and therapy, Nature Reviews Clinical Oncology 15 (7) (2018) 409–421. doi:10.1038/s41571-018-0018-y. URL https://www.nature.com/articles/s41571-018-0018-y

[10] E. Terpos, L. A. Moulopoulos, M. A. Dimopoulos, Advances in imaging and the management of myeloma bone disease, Journal of Clinical Oncology: Official Journal of the American Society of Clinical Oncology 29 (14) (2011) 1907–1915. doi:10.1200/JCO.2010.32.5449.

[11] J. Guinea-Pérez, A. Ceresi, A. F. García, B. A. Galende, A. Belmonte-Hernández, S. Peluso, F. Alvarez, Radiomics feature analysis for survival prediction in multiple myeloma: An automated PET/CT approach, Computer Methods and Programs in Biomedicine 271 (2025) 109019. doi:10.1016/j.cmpb.2025.109019. URL https://www.sciencedirect.com/science/article/pii/ S0169260725004365

[12] J. Shreve, C. Charalampous, R. Warsame, J. Pritchett, P. Kapoor, F. Buadi, J. Paludo, D. Dingli, M. Shah, A. Fonder, S. Hayman, N. Leung, M. Gertz, R. Kyle, S. Broski, V. Rajkumar, S. Kumar, M. Binder, P921: ARTIFICIAL INTELLIGENCE BASED FDG PET/CT RADIOMICS FOR RISK STRATIFICATION IN NEWLY DIAGNOSED MULTIPLE MYELOMA, HemaSphere 7 (Suppl) (2023) e156011e. doi: 10.1097/01.HS9.0000970588.15601.1e. URL https://www.ncbi.nlm.nih.gov/pmc/articles/PMC10431033/

[13] H. Zhong, D. Huang, J. Wu, X. Chen, Y. Chen, C. Huang, 18F-FDG PET/CT based radiomics features improve prediction of prognosis: multiple machine learning algorithms and multimodality applications for multiple myeloma, BMC Medical Imaging 23 (1) (2023) 87. doi: 10.1186/s12880-023-01033-2.

[14] B. Ni, G. Huang, H. Huang, T. Wang, X. Han, L. Shen, Y. Chen, J. Hou, Machine Learning Model Based on Optimized Radiomics Feature from 18F-FDG-PET/CT and Clinical Characteristics Predicts Prognosis of Multiple Myeloma: A Preliminary Study, Journal of Clinical Medicine 12 (6) (2023) 2280. doi:10.3390/jcm12062280.

[15] J. L. Katzman, U. Shaham, A. Cloninger, J. Bates, T. Jiang, Y. Kluger, DeepSurv: personalized treatment recommender system using a Cox proportional hazards deep neural network, BMC Medical Research Methodology 18 (1) (2018) 24. doi:10.1186/s12874-018-0482-1.

[16] C. Lee, W. Zame, J. Yoon, M. v. d. Schaar, DeepHit: A Deep Learning Approach to Survival Analysis With Competing Risks, Proceedings of the AAAI Conference on Artificial Intelligence 32 (1), number: 1 (Apr. 2018). doi:10.1609/aaai.v32i1.11842. URL https://ojs.aaai.org/index.php/AAAI/article/view/11842

[17] S. Wiegrebe, P. Kopper, R. Sonabend, B. Bischl, A. Bender, Deep learning for survival analysis: a review, Artificial Intelligence Review 57 (3) (2024) 65. doi:10.1007/s10462-023-10681-3.

[18] R. Bommasani, D. A. Hudson, E. Adeli, R. Altman, S. Arora, S. v. Arx, M. S. Bernstein, J. Bohg, A. Bosselut, E. Brunskill, E. Brynjolfsson, S. Buch, D. Card, R. Castellon, N. Chatterji, A. Chen, K. Creel, J. Q. Davis, D. Demszky, C. Donahue, M. Doumbouya, E. Durmus, S. Ermon, J. Etchemendy, K. Ethayarajh, L. Fei-Fei, C. Finn, T. Gale, L. Gillespie, K. Goel, N. Goodman, S. Grossman, N. Guha, T. Hashimoto, P. Henderson, J. Hewitt, D. E. Ho, J. Hong, K. Hsu, J. Huang, T. Icard, S. Jain, D. Jurafsky, P. Kalluri, S. Karamcheti, G. Keeling, F. Khani, O. Khattab, P. W. Koh, M. Krass, R. Krishna, R. Kuditipudi, A. Kumar, F. Ladhak, M. Lee, T. Lee, J. Leskovec, I. Levent, X. L. Li, X. Li, T. Ma, A. Malik, C. D. Manning, S. Mirchandani, E. Mitchell, Z. Munyikwa, S. Nair, A. Narayan, D. Narayanan, B. Newman, A. Nie, J. C. Niebles, H. Nilforoshan, J. Nyarko, G. Ogut, L. Orr, I. Papadimitriou, J. S. Park, C. Piech, E. Portelance, C. Potts, A. Raghunathan, R. Reich, H. Ren, F. Rong, Y. Roohani, C. Ruiz, J. Ryan, C. Ré, D. Sadigh, S. Sagawa, K. Santhanam, A. Shih, K. Srinivasan, A. Tamkin, R. Taori, A. W. Thomas, F. Tramèr, R. E. Wang, W. Wang, B. Wu, J. Wu, Y. Wu, S. M. Xie, M. Yasunaga, J. You, M. Zaharia, M. Zhang, T. Zhang, X. Zhang, Y. Zhang, L. Zheng, K. Zhou, P. Liang, On the Opportuni-ties and Risks of Foundation Models, arXiv:2108.07258 [cs] (Jul. 2022). doi:10.48550/arXiv.2108.07258. URL http://arxiv.org/abs/2108.07258

[19] A. Radford, J. W. Kim, C. Hallacy, A. Ramesh, G. Goh, S. Agarwal, G. Sastry, A. Askell, P. Mishkin, J. Clark, G. Krueger, I. Sutskever, Learning Transferable Visual Models From Natural Language Supervision, arXiv:2103.00020 [cs] (Feb. 2021). doi:10.48550/arXiv.2103.00020. URL http://arxiv.org/abs/2103.00020

[20] A. Kirillov, E. Mintun, N. Ravi, H. Mao, C. Rolland, L. Gustafson, T. Xiao, S. Whitehead, A. C. Berg, W.-Y. Lo, P. Dollár, R. Girshick, Segment Anything, arXiv:2304.02643 [cs] (Apr. 2023). doi:10.48550/arXiv.2304.02643. URL http://arxiv.org/abs/2304.02643

[21] W. Khan, S. Leem, K. B. See, J. K. Wong, S. Zhang, R. Fang, A Com-prehensive Survey of Foundation Models in Medicine, arXiv:2406.10729 [cs] (Jan. 2025). doi:10.48550/arXiv.2406.10729. URL http://arxiv.org/abs/2406.10729

[22] J. Ma, Y. He, F. Li, L. Han, C. You, B. Wang, Segment anything in medical images, Nature Communications 15 (1) (2024) 654. doi:10.1038/s41467-024-44824-z. URL https://www.nature.com/articles/s41467-024-44824-z

[23] J. Ma, S. Kim, F. Li, M. Baharoon, R. Asakereh, H. Lyu, B. Wang, Segment Anything in Medical Images and Videos: Benchmark and De-ployment, arXiv:2408.03322 [eess] (Aug. 2024). doi:10.48550/arXiv.2408.03322. URL http://arxiv.org/abs/2408.03322

[24] W. Zhu, Y. Chen, S. Nie, H. Yang, SAMMS: Multi-modality Deep Learning with the Foundation Model for the Prediction of Cancer Patient Survival, in: 2023 IEEE International Conference on Bioinformatics and Biomedicine (BIBM), 2023, pp. 3662–3668. doi:10.1109/BIBM58861.2023.10385661. URL https://ieeexplore.ieee.org/document/10385661

[25] A. Palumbo, H. Avet-Loiseau, S. Oliva, H. M. Lokhorst, H. Goldschmidt, L. Rosinol, P. Richardson, S. Caltagirone, J. J. Lahuerta, T. Facon, S. Bringhen, F. Gay, M. Attal, R. Passera, A. Spencer, M. Offidani, S. Kumar, P. Musto, S. Lonial, M. T. Petrucci, R. Z. Orlowski, E. Zamagni, G. Morgan, M. A. Dimopoulos, B. G. M. Durie, K. C. Anderson, P. Sonneveld, J. San Miguel, M. Cavo, S. V. Rajkumar, P. Moreau, Revised International Staging System for Multiple Myeloma: A Report From International Myeloma Working Group, Journal of Clinical Oncology: Official Journal of the American Society of Clinical Oncology 33 (26) (2015) 2863–2869. doi:10.1200/JCO.2015.61.2267.

[26] L. K. S. Sundar, J. Yu, O. Muzik, O. C. Kulterer, B. Fueger, D. Kifjak, T. Nakuz, H. M. Shin, A. K. Sima, D. Kitzmantl, R. D. Badawi, L. Nardo, S. R. Cherry, B. A. Spencer, M. Hacker, T. Beyer, Fully Automated, Semantic Segmentation of Whole-Body ^18^F-FDG PET/CT Images Based on Data-Centric Artificial Intelligence, Journal of Nuclear Medicine 63 (12) (2022) 1941. doi:10.2967/jnumed.122.264063. URL http://jnm.snmjournals.org/content/63/12/1941.abstract

27. [27] J. Ma, Z. Yang, S. Kim, B. Chen, M. Baharoon, A. Fallahpour, R. Asakereh, H. Lyu, B. Wang, MedSAM2: Segment Anything in 3D Medical Images and Videos, arXiv:2504.03600 [eess] (Apr. 2025). doi:10.48550/arXiv.2504.03600. URL http://arxiv.org/abs/2504.03600

[28] N. Ravi, V. Gabeur, Y.-T. Hu, R. Hu, C. Ryali, T. Ma, H. Khedr, R. Rädle, C. Rolland, L. Gustafson, E. Mintun, J. Pan, K. V. Alwala, N. Carion, C.-Y. Wu, R. Girshick, P. Dollár, C. Feichtenhofer, SAM 2: Segment Anything in Images and Videos, arXiv:2408.00714 [cs] (Oct. 2024). doi:10.48550/arXiv.2408.00714. URL http://arxiv.org/abs/2408.00714

[29] J. Hu, L. Shen, S. Albanie, G. Sun, E. Wu, Squeeze-and-Excitation Networks, arXiv:1709.01507 [cs] (May 2019). doi:10.48550/arXiv.1709.01507. URL http://arxiv.org/abs/1709.01507

[30] M. Claesen, J. Simm, D. Popovic, Y. Moreau, B. D. Moor, Easy Hyperparameter Search Using Optunity, arXiv:1412.1114 (Dec. 2014). doi:10.48550/arXiv.1412.1114. URL http://arxiv.org/abs/1412.1114

[31] G. C. Cawley, N. L. C. Talbot, On Over-fitting in Model Selection and Subsequent Selection Bias in Performance Evaluation.

[32] F. E. Harrell, R. M. Califf, D. B. Pryor, K. L. Lee, R. A. Rosati, Evaluating the yield of medical tests, JAMA 247 (18) (1982) 2543–2546.

[33] K. He, X. Zhang, S. Ren, J. Sun, Deep Residual Learning for Image Recognition, 2016, pp. 770–778. URL https://openaccess.thecvf.com/content_cvpr_2016/html/He_Deep_Residual_Learning_CVPR_2016_paper.html

[34] Updates to Management of Multiple Myeloma, Journal of the National Comprehensive Cancer Network 23 (Supplement) (May 2025). doi:10.6004/jnccn.2025.5008. URL https://jnccn.org/view/journals/jnccn/23/Supplement/ article-e255008.xml

[35] E. Zamagni, C. Nanni, F. Gay, A. Pezzi, F. Patriarca, M. Bellò, I. Rambaldi, P. Tacchetti, J. Hillengass, B. Gamberi, L. Pantani, V. Magarotto, A. Versari, M. Offidani, B. Zannetti, F. Carobolante, M. Balma, P. Musto, M. Rensi, K. Mancuso, A. Dimitrakopoulou-Strauss, S. Chauviè, S. Rocchi, N. Fard, G. Marzocchi, G. Storto, P. Ghedini, A. Palumbo, S. Fanti, M. Cavo, 18F-FDG PET/CT focal, but not osteolytic, lesions predict the progression of smoldering myeloma to active disease, Leukemia 30 (2) (2016) 417–422. doi: 10.1038/leu.2015.291. URL https://www.nature.com/articles/leu2015291

[36] C. Ryali, Y.-T. Hu, D. Bolya, C. Wei, H. Fan, P.-Y. Huang, V. Aggarwal, A. Chowdhury, O. Poursaeed, J. Hoffman, J. Malik, Y. Li, C. Feichtenhofer, Hiera: a hierarchical vision transformer without the bells-and-whistles, in: Proceedings of the 40th International Conference on Machine Learning, Vol. 202 of ICML’23, JMLR.org, Honolulu, Hawaii, USA, 2023, pp. 29441–29454.

[37] K. Clark, B. Vendt, K. Smith, J. Freymann, J. Kirby, P. Koppel, S. Moore, S. Phillips, D. Maffitt, M. Pringle, L. Tarbox, F. Prior, The Cancer Imaging Archive (TCIA): maintaining and operating a public information repository, Journal of Digital Imaging 26 (6) (2013) 1045–1057. doi:10.1007/s10278-013-9622-7.

